# Personal Resources, Emotion Regulation, Parenting Style, Social Support On Cyberbullying Victims: A Literature Review

**DOI:** 10.1101/2021.12.09.21267506

**Authors:** Nia Agustiningsih, Moses Glorino Rumambo Pandin

## Abstract

Cyberbullying is a teenage phenomenon that needs attention because it causes serious short-term and long-term impacts in all aspects of health. The right coping strategies are needed by teenagers to be able to face cyberbullying so that teenagers have good resilience. The purpose of this literature review is to find out what are the important things that need to be considered in cyberbullying victims. The method of writing this article is a literature review of 25 articles with the year 2019-2021 published from the Scopus, Science direct, SAGE and Proquest databases. The method of searching and selecting articles used *The Center for Review and Dissemination and the Joanna Briggs Institute Guideline* and PRISMA *Checklist* with selection criteria using the PICOS approach. The results of the literature review show that Personal Resources, Emotional Regulation, Parenting Patterns and Support from Peers, Family/Parents, and Teachers is an important thing that needs to be considered for victims of cyberbullying. Furthermore, Adolescents need to get promotive and interventional programs to improve their abilities in coping strategies against cyberbullying and bullying so that with the right coping strategies they will feel good welfare and optimal academic achievement at school.

## INTRODUCTION

Bullying is violent behavior or attacks in the form of verbal or non-verbal (physical) aimed at the victim by the perpetrator which causes psychological, emotional and physical impacts because the victim is unable to defend against attacks that are carried out continuously which can affect the victim in several aspects such as social, emotional, health and academic[1]. The term cyberbullying is related to bullying that is carried out via the internet / online using information and communication technology. Compared to traditional bullying, cyberbullying reaches an unlimited number of people with increased exposure and time[10]. Cyberbullying is bullying that is carried out using an internet network system such as sending rude messages, mocking, hurting or embarrassing and harassing images through social media), intimidating via telephone calls and distributing videos that harass the victim, posting disparaging comments and/or embarrassing images on the social networks, or to intimidate or threaten someone electronically[9,15].

Cyberbullying is recognized as a serious growing psychosocial problem occurring in schools around the world[18,21]. Most of the studies on the prevalence of cyberbullying have been carried out based on different age groups of students from primary, secondary school. The results of the school population show that cyberbullying victims vary from 10% to 53% which describes the phenomenon in all age groups of students[27]. Cyberbullying has a significant negative impact on emotion well being in adolescents which is shown as negative feelings and emotions such as stress, sorrowful, angry, frustrated, shame, lonely, fright, heaviness, desire for revenge and suicidal idea[4,12,19,25]. Cyberbullying also has an impact on behavior problems such as juvenile delinquency, violence and bad value shift and functions at school until dropping out and withdrawing from the social environment[20]. This is a digital era, so every teenager will be at risk of cyberbullying, so teenagers need to know the right coping strategies to deal with this phenomenon[8]. In addition to the coping strategies used against cyberbullying, other strategies are needed that are different from traditional bullying[3].

Coping strategies are efforts made by individuals to adapt to stress as a result of cyberbullying so that they have resistance to psychological disorders. Emotion focused coping is problem solving performed to manage negative feelings resulting from a stressful event. Problem focused coping centers on solving problems that occur as a result of events. Various types of problem focused coping strategies used to deal with cyberbullying such as blocking the cyberbully’s contact and direct confrontation are the most relevant strategies [3]. The results showed that victims who had direct confrontation stated that they were effective in stopping bullying. By doing confrontation, adolescents learn to recognize stressful situations as an effort to grow and develop in having resilience[3]. In dealing with cyberbullying, adolescents need to be prepared with education and training related to the selection of coping strategies through education about emotional intelligence, emotional regulation, emotional social competence as described in the paragraph above so that adolescents have resilience in the face of cyberbullying. Based on this description, the systematic writing of this literature review aims to determine the perspective of the factors that play a role in the selection of adolescent coping strategies to face cyberbullying.

## METHOD

The approach used in this activity is a literature review. The method of implementing the activities carried out is the process of collecting data:

1. Prepare a literature review scientific paper design according to a summary of the topic to be carried out.
2. Determine and prepare the registration protocol using the *Boolean* operator (AND, OR NOT or AND NOT). Keywords in this literature review are adjusted to Medical Subject Heading (MeSH).
  a. Determining which *database* to use and which to use in this activity is to use *Scopus, ProQuest, Science Direct, SAGE*, with the article year 2019 – 2021.
  b. Determine the eligibility criteria with an article search strategy using the PICOS framework which is also adjusted to the inclusion and exclusion criteria (table 1)
  c. Explain the source of information in searching articles in a predetermined database until the last article is found for a comprehensive summary
  d. The study selection process by reading the entire article and selecting the articles that did not fit was discarded and recorded in the selection strategy using the PRISMA flow chart (figure 1). The article analysis method used in this literature review is a comprehensive process consisting of four interrelated stages: Reading the contents of the article in depth from all research results, Marking line by line the study findings, compiling sentences or findings marked based on similarity of meaning into themes presented in descriptive writing, and interpreting the analytical themes then written in the results of the *review*. In addition, descriptive analysis in the form of narrative is also added

**Table 1.** Inclusion and exclution Criteria with PICOS

| Criteria | Inclusion | Exclusion |
| --- | --- | --- |
| **Population** | Adolescence | Non adolescence age |
| **Intervention** | There are no intervention | There are no criteria exclusion |
| **Comparisons** | There are no comparison | There are no criteria exclusion |
| **Outcome** | Strategies coping to cyberbullying | No relevant Strategies coping to *cyberbullying* |
| **Study Type** | Cross sectional, longitudinal study | mixed method, a quasi experimental design, clinical trial or randomized controlled trial, Systematic or literature reviews, qualitative research |
| **Publication Type** | Peer reviewed original studies | Non peer-reviewed studies |
| **Publication Years** | 2019 – 2021 | Pre 2019 |
| **Language** | English | Language ather than English |

**Figure 1.**
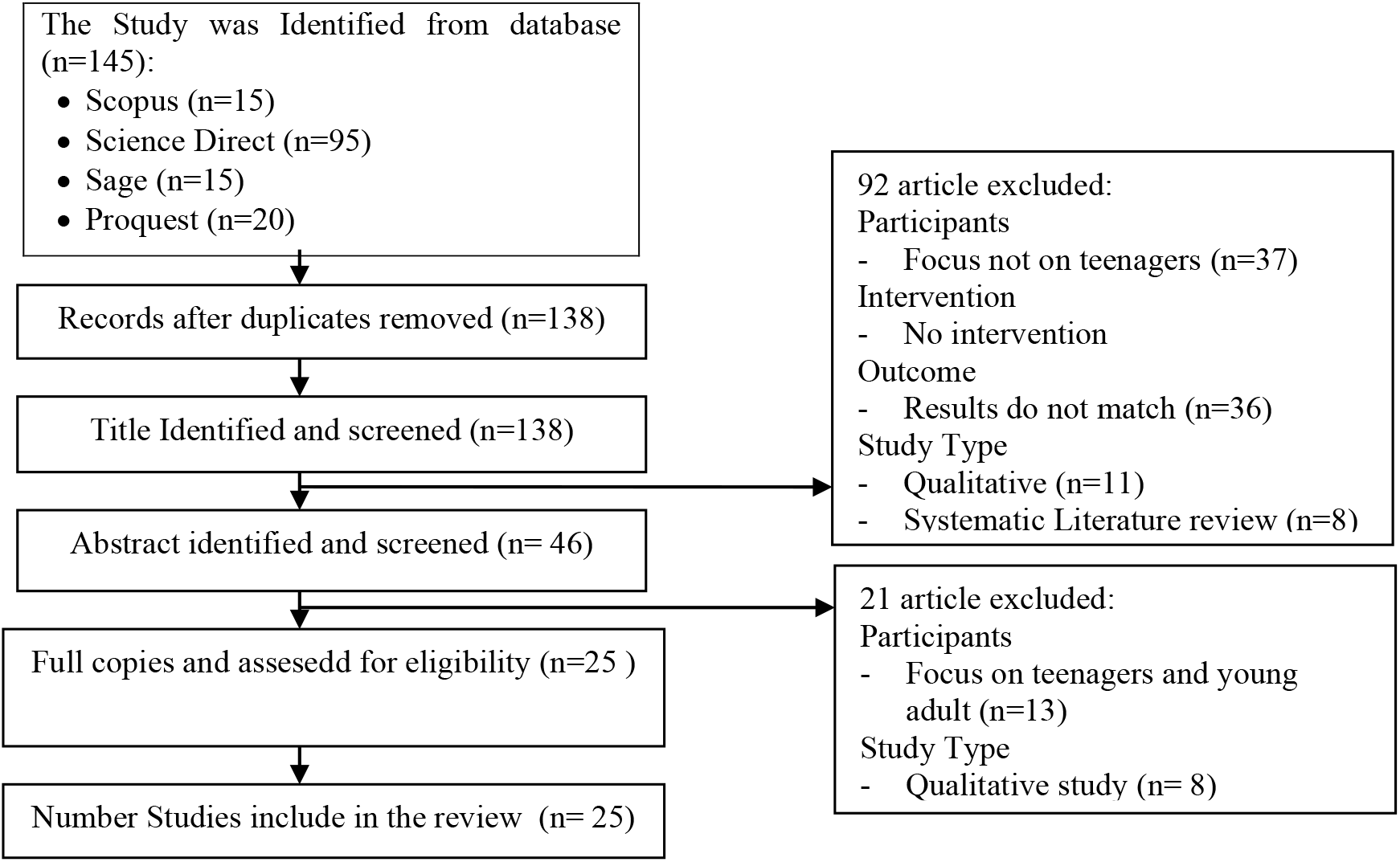
PRISMA Flow Chart

## RESULTS

The results of the study of 25 articles obtained the following things:

### 1. Personal Trait

Adolescents who feel lonely have a history of anxiety, suicidal ideation and attempts. Age, loneliness, anxiety, suicidal ideation and attempts, and marijuana use are risk factors for bullying victims. Education about behavior and emotional expression is needed to strengthen students’ coping with bullying. Cyberbullying victimization is positively associated with social anxiety and depression [1,23]. It is very important to improve children’s social skills and modify negative cognitions in intervention programs to help children receive feedback on other people’s reactions. Differences in anxiety, emotional dependence, social support, and social skills between direct actors, control actors, and direct/control actors [22]. Intervention programs are needed for the development of social interpersonal and individual cognitive factors. Cyberbullying victimization is associated with nonsuicidal self-injury (NSSI) through increased depression and early maladaptive schemas (EMS) with mindfulness as a protective factor. Cyberbullying, mindfulness and depression are interrelated [24]. It is necessary to improve intervention programs for adolescents through intervention programs that involve mindfulness as an effort to increase resilience or as a protective factor for adolescents in the face of cyberbullying [7]. There is a need for promotive efforts related to regulating positive emotions such as mindfulness attention and strategies for reducing emotion regulation for negative emotions such as acceptance, seeking support. Cyberbullying victims who showed symptoms of depression reported lower levels of personal resources (emotional intelligence, gratitude, optimism, and forgiveness) than those who did not [17]. The importance of understanding through promotional and preventive programs relates to personal resources in mental health and in helping individuals to have resilience in the face of stressors such as cyberbullying.

### 2. Moral Guidance

Students with positive/loving emotions about bullying increase their active behavior in dealing with bullying by re-offending bullying [4]. Students with negative emotions/do not like bullying increase proactive behavior in dealing with bullying such as seeking support from adults and telling them to stop bullying. Students with high moral disengagement are associated with passive behavior when facing bullying while students with low moral disengagement are associated with proactive behavior in dealing with bullying such as seeking support from adults and telling them to stop bullying.Teenagers need to get moral guidance so that they have the awareness to stop bullying which is a harmful act. In a study conducted on 274 adolescents, it was found that confrontation with cyberbullying is negatively related to emotional symptoms [3]. Coping strategies in dealing with bullying and cyberbullying develop according to age. Parenting style is related to adolescent involvement in bullying/cyberbullying behavior [14]. The importance of knowing the values that must be socialized by parents to children because this is related to the involvement of adolescents in bullying and cyberbullying. Moral emotions mediate the relationship between parenting and cyberbullying. The importance of nurturing and providing moral values so that it can grow moral norms in children [25].

### 3. Emotional understanding and regulation

Adolescents who have a high level of emotional understanding and regulation of emotions have a lower likelihood of becoming victims, victims and perpetrators of cyberbullying [6]. The need for intervention programs and prevention education for adolescents in dealing with cyberbullying actions in adolescents by increasing the ability of Emotional intelligence. Adolescents with higher levels of Difficulties in Emotion Regulation had high levels of loneliness, which also led to higher levels of depression, and ultimately were at risk for cyberbullying aggression [10]. It is necessary to increase the ability of emotion regulation in adolescents as a strategy for adolescents in dealing with cyberbullying so that they are able to manage emotions and not take revenge or cyberbullying aggression.Gratitude gratitude is an intermediary in the relationship between emotional intelligence and bullying attacks through cyberspace [5]. Adolescents need to acquire emotional and educational skills to increase gratitude as a strategy for dealing with cyberbullying. Education about improving psychological well-being is important for adolescents which aims to strengthen affective bonds and emotional self-regulation strategies as the main source, without ignoring the role of social cognition and perceptions of social support. Emotional Intelligence is positively related to satisfaction with life and negatively related to bullying and cyberbullying in adolescents. Control variables such as gender, age, class, self-emotion appraisal, use of emotions and emotion regulation are good predictors of life satisfaction for victims of bullying and cyberbullying [15]. Emotional intelligence needs to be promoted as an effort to improve the welfare of adolescents who are involved in bullying and cyberbullying. Furthermore, Intervention is needed for adolescent victims of cyberbullying to develop affective abilities so that they can carry out adaptive emotional regulation to obtain a good degree of health through intervention programs. Teenagers who are victims of bullying who have low levels of emotional intelligence and flourishing are associated with high suicide risk, on the other hand, suicide risk is lower in adolescents who have higher emotional intelligence [18]. The importance of understanding the role of Emotional intelligence and needs to be developed in preventing the risk of suicide among adolescents who are victims of cyberbullying.

### 4. Social support

Teenagers face cyberbullying by telling their parents, teachers, or friends [4,8,22]. Only a few teens directly address the problem by asking the cyberbully to stop texting or calling the abuser and, in some cases, the problem is solved by turning off their cell phones. Strategies for preventing and managing cyberbullying must be considered as part of the overall educational approach so that psychoeducation is needed for adolescents to find out strategies for dealing with cyberbullying, besides that a policy is needed to introduce cyberbullying to the curriculum and conduct training for teachers and education for parents. Family support is important for cyberbullying victims[13]. Education about cyberbullying in families is very important.

### 5. Communication and decision making skills

Most of the individual factors are significantly related to Traditional Victims, Cyber Victims [9]. It is very important to develop and improve problem-solving communication skills, and decision-making skills to increase assertiveness in dealing with traditional bullying and cyberbullying. Most students in dealing with cyberbullying prefer to remain silent because they feel they are not accepted in the group or even some are removed or removed from the group[11]. Improving personal skills through communication skills and emotional regulation is important for adolescents. Socioemotional and relationship skill related to being victims of bullying and cyberbullying [12]. Good relationship skills which are part of socio-emotional competencies need to be developed for all genders in school-age children and adolescents as an effort for teenagers to be able to face bullying and cyberbullying. Age is positively related to the use of problem-solving coping skills to regulate behavior against cyberbullying [19]. Problem-solving coping skills need to be developed in adolescents to face cyberbullying. Cyberbullying victimization is positively associated with social anxiety and depression [23]. It is very important to improve children’s social skills and modify negative cognitions in intervention programs to help children receive feedback on other people’s reactions.

## DISCUSSION

Coping strategies that can be done by adolescents in dealing with cyberbullying are reacting directly (such as retaliating), neglecting (avoiding), getting support (parents, friends, or teachers), and blocking message[14]. Another coping strategy is technical coping, which is the most frequently used coping strategy followed by assertiveness, support from close people, helplessness/self-blame, retaliation, and advice[8]. Teenagers need to receive regular training to acquire proficiency to deal with cyberbullying, help other teenagers avoid from revenge motivation. Active strategies for dealing with cyberbullying are effective for dealing with problems related to cyberbullying[14]. Although problem-solving techniques can reduce the psychological impact of cyberbullying, an estimated 25% of cyberbullying victims do not seek support, and only one in three teens tell their parents about cyberbullying[20]. The role selection of adolescent coping strategies to on Cyberbullying:

### 1. Personal Resources

Efforts in dealing with cyberbullying are determined by each individual’s personal resources such as emotional intelligence, gratitude and cognitive assessment and mindfulness[5,24,26]. Emotional intelligence is a mental ability that functions to understand, identify and manage/regulate one’s own emotions and act with others[7]. Gratitude is a protective factor that can improve adolescent mental health in dealing with cyberbullying[5,7]. In general, better emotional management skills are associated with life satisfaction and better psychological adjustment. In general, better emotional management skills are associated with life satisfaction and better psychological adjustment. The results showed that adolescents with high emotional understanding were able to cope with stress better, while adolescents with low levels of emotional regulation showed social anxiety and more symptoms of stress, low self-esteem, depression, low life satisfaction, risk of suicide [5,16,19,23]. Emotional intelligence component consists of the capability to assess one’s own, others, use and regulate emotions[16].

### 2. Emotion regulation

Emotion regulation is the competency to manage emotions socially[11]. Emotion regulation consists of the ability to perceive, understand[17], receive emotional responses, and avoid impulsive behavior when angry, and engage in effective emotion regulation strategies. According to the emotion regulation process model, effective emotion regulation can reduce ongoing negative emotion expressive[2,22]. In this case emotion regulation is also related to socio-emotional competence which includes emotional skills, understanding and attitudes as well as social competence. These include responsibility in making decisions, skills in building relationships with others, self and social awareness[13]. Proactive behavior and knowledge about steps to deal with cyberbullying/intimidation are important for teenagers[4]. These skills are useful for socio-emotional development, friendship formation, and behavioral socialization in children and adolescents in establishing relationships with peers[25].

### 3. Parenting Style

Parenting patterns are related to the involvement of children in bullying and or cyberbullying both as perpetrators, victims, victims - perpetrators. In parenting, parents need to know the values that must be instilled by parents in children because this is related to the involvement of teenagers in bullying and cyberbullying such as socialization or the introduction and inculcation of values and morals[15]. Socialization on moral development is beneficial throughout childhood, adolescence and adulthood which consists of empathy, courage and moral identity[6]. A person’s moral identity is stored in memory as a complex knowledge structure consisting of moral values, goals, traits, and behavior. A person’s morality and sense of identity are intertwined.

### 4. Peer-support, Family or Parents, and Teachers

Adolescents who have support are less likely to be victims of intimidation than those who do not. Involvement of peers as educators (helping to awaken) can reduce the victimization of bullying in adolescents[9]. Therefore, increasing peer awareness, empathy and self-efficacy to support fellow victims can be part of an anti-bullying program in schools[14]. Individuals who receive strong parental support and get parental control that is oriented towards affection in internalizing moral norms will help adolescents in dealing with cyberbullying/bullying[14]. Parental support makes adolescents have the ability to develop resilience in the face of cyberbullying/bullying such as by trying to do confrontation, increasing personal agency and individual trust[4].

Teacher support is very necessary for teenagers in dealing with cyberbullying/bullying because teachers are an important component in the school environment that is able to provide comfort to their students. Several studies explain that adolescents who feel an uncomfortable environment at school will tend to avoid cyberbullying and bullying by not attending school because they feel there is no source of support, including teachers[14]. Therefore, training or socialization for teachers is needed as an important component in schools related to training programs to deal with cyberbullying and bullying for both perpetrators, victims, perpetrators - victims.

## CONCLUSION

Adolescents need to get promotive and interventional programs to improve their abilities in coping strategies against cyberbullying and bullying so that with the right coping strategies they will feel good welfare and optimal academic achievement at school.

## Data Availability

All data produced in the present work are contained in the manuscript

